# Accelerated neurodevelopment of reward anticipation processing in adolescent girls with depression

**DOI:** 10.1101/2023.09.15.23295631

**Authors:** David AA Baranger, Morgan Lindenmuth, Leehyun Yoon, Amanda E. Guyer, Kate Keenan, Alison E Hipwell, Erika E Forbes

## Abstract

**Objective:** To test the hypothesis that depression is associated with differential neurodevelopment of reward circuitry in adolescence.

**Methods:** Adolescent girls (N=183, 58 with MDD in early or late adolescence) underwent MRI scans from ages 16-20 (1-4 scans/participant, 477 scans total) and completed a card-guessing fMRI task with monetary rewards. Mixed-effect models tested the effect of age and the moderating effect of MDD on whole-brain regional activation during reward anticipation.

**Results:** Eighty of 414 regions showed age effects (p_FDR_<0.05), consisting primarily of increasing activation with increasing age. Most significant regions were in dorsal attention, salience, and somatomotor networks, and also included the bilateral putamen, pallidum, and right nucleus accumbens. MDD moderated age effects in 40 regions (p_FDR_<0.05), including the right putamen, medial orbitofrontal cortex, and amygdala, and regions in control and dorsal attention networks. MDD x linear and quadratic age effects were negative, suggesting that MDD was associated with accelerated neurodevelopment.

**Conclusions:** Theories of reward processing’s contribution to adolescent risk for depression focus primarily on core reward regions, yet a host of regions beyond these continue to develop during late adolescence. Findings demonstrate differing regional patterns of age-related changes in relation to MDD in girls, suggesting that depression involves disruption of a wide range of regions during reward anticipation processing across adolescence. Childhood and adolescent MDD is associated with accelerated neurodevelopment of attention and cognitive control regions during reward anticipation processing, which may have consequences both for cognitive function and the emergence of reward-system-specific disruptions.

## Introduction

The hallmark symptoms of depression include sadness, fatigue and anhedonia, which have been hypothesized to reflect altered neural processing of rewards^1^. Blunted activation of the ventral striatum (VS) during reward processing has emerged as a highly consistent finding in meta-analyses of major depressive disorder (MDD)^2, 3^. Moreover, reward-related activation prospectively predicts MDD and depression symptoms^4–6^, is altered in youth with familial risk for depression^7–10^, and predicts treatment response in MDD^11, 12^. Collectively, these findings suggest a potential mechanistic role of reward processing in MDD^13^, which is further supported by observations that reward circuitry manipulations influence depressive-like behaviors in model organisms^14, 15^.

The peak incidence for MDD occurs during adolescence^16–18^, especially for girls^19^. Adolescence is also a developmental period in which reward-related activation, particularly of the VS, changes and peaks^20–22^. Evidence suggests that adolescents with MDD show greater VS blunting than adults with MDD^2, 8^, and there are sex differences in the maturational time-course of reward-related brain structures^23^ and associated behaviors^24, 25^. Thus, one emerging hypothesis is that differences in the maturational time course of reward processing in the VS^1, 21, 26^ contributes to depression beyond differences in reward function in depressed relative to non-depressed participants. Such developmental differences may be most evident in higher-risk populations, such as adolescent girls, and may even partially underlie the elevated incidence of depression in adolescent girls.

One proposed developmental model is that VS activation is chronically low in MDD--blunted activation in adolescents with MDD is a product of a smaller developmentally expected increase in reward activity^1, 8^. An alternative model is that adolescents with, or at risk for, MDD display accelerated neurodevelopment, such that reward processing in the VS peaks and decreases earlier relative to non-depressed adolescents—blunted activation is thus a by-product of being at a later neurodevelopmental point^5, 27, 28^. Adjudicating between these models requires testing whether, and how, the correlation between age and reward processing differs in MDD. To date, findings have been limited, as age is largely treated as a confound in neuroimaging studies of MDD, and few studies of adolescent depression have measured activation longitudinally. Understanding the nature of developmental disruption in reward processing in adolescent girls with depression is critical to elucidating the etiology and pathophysiology of this form of psychopathology.

Prior empirical and theoretical literature has focused primarily on the VS, driven both by its central role in reward processing^29^ and by clear associations with MDD^2, 3^. However, evidence indicates that regions throughout the brain play critical roles in reward processing^29, 30^ and reward signals are present in networks that play primary roles in other functions (including vision and attention)^31, 32^. This focus on a subset of regions may have led the field to miss other important associations^33^. Indeed, in a recent meta-analysis of reward processing in MDD, 15 of 38 included studies examined associations only within *a priori* regions of interest^2^. Additionally, the majority of studies examining reward processing during development have compared adolescents and adults, rather than tracking neurodevelopment through adolescence. Thus, knowledge on the time-course of reward processing development during adolescence is limited^34–36^, with the exception of the VS^20^. These gaps highlight the need for studies that examine whole-brain reward processing changes during adolescence and that test whether the development of reward processing activation differs in individuals with a history of MDD.

The present study examined the development of whole-brain activation during reward anticipation in a sample of adolescent girls scanned longitudinally from ages 16-20. Analyses focused on reward anticipation, given meta-analytic evidence that the correlation of MDD with striatal activation to reward anticipation, but not reward receipt, may vary with age^2^ and that striatal reward anticipation activation is predictive of future changes in depressive symptoms^13^. Analyses sought first to test whether whole-brain reward anticipation activation changes with age during late adolescence and second to test whether group-level trajectories differed between participants with and without childhood or adolescent MDD, controlling for other known correlates of reward processing and depression, including pubertal maturation^20^, financial strain^37^, and race^38^. It was hypothesized that curvilinear patterns of activation would be observed across development^20, 36^, particularly in canonical reward regions (e.g., VS). Based on evidence of accelerated development of brain structure^27^ and accelerated cellular aging in MDD^39^, it was hypothesized that participants with MDD would show a pattern of neurodevelopment consistent with accelerated maturation of reward-related activation in late adolescence.

## Methods

### Sample

Participants were N=183 girls, ages 16-20. Participants and their mothers were recruited from the longitudinal Pittsburgh Girls Study (PGS)^40^, which followed an enumeration of households including girls between the ages of 5 and 8 in the city of Pittsburgh. Of 2990 eligible families, 2450 (85%) were successfully re-contacted and agreed to participate in a prospective study. A subset of PGS participants was recruited to the PGS Emotions sub-study (PGS-E), a study of precursors to depression (*N*=232^41^), at age 9. Eight year old PGS participants who scored in the top 25% on self-and/or maternal report of depression and a randomly selected group of girls scoring in the lower 75% (matched on race) were targeted for enrollment in the PGS-E. Participants identified as Black/African American (*n*=124, 68%), White (*n*=50, 27%), or multi-racial (*n*=9, 5%). Participants completed annual diagnostic interviews from ages 9-13 years. From ages 16-20, participants completed up to four annual study visits and fMRI scanning sessions. All procedures received Institutional Review Board approval at the University of Pittsburgh and all participants (when they reached age 18 years) and their mothers provided informed consent for their participation. Of the potential *k*=928 scans (4 per participant, *N*=232 participants), the final sample consisted of k=477 reward fMRI scans from *N*=183 participants pooled across the four annual assessments. Scans were unavailable or excluded from analyses because participants (1) withdrew or could not be scheduled (*k*=228 scans, *n*=121 participants), (2) were ineligible for an MRI scan due to medical or physical exclusionary criteria (k=80 scans, *n*=55 participants), (3) refused or were unable to complete the MRI portion of the study (k=69, *n*=48 participants), (4) did not correctly perform the reward task or scans did not pass MRI quality control benchmarks (*k=*86, *n*=65 participants). In the final sample, 49 participants had four scans, 51 had three, 45 had two, and 38 had one (Supplemental Figure 1). The average time between scans was 1.27 years (SD=0.5; range=0.44-4.18).

### Assessment of Depression

The Kiddie-Schedule for Affective Disorders and Schizophrenia-Present and Lifetime Version (KSADS-PL)^42^ was administered at every study visit. The KSADS assesses both child-reported and caregiver-reported depression symptoms through age 18 years, at which time report by caregivers was discontinued. At ages 9 to 12 girls and caregivers were asked about depression symptoms in the past month, and at ages 13 and older were asked about symptoms in the past month and past year. Participants were classified as having had MDD if they met DSM-IV criteria at any study visit by combined parent and child report (n=58; n=31 met criteria at ages 9-14; n=27 met criteria at ages 16-20). Participants with MDD did not differ from those without MDD on sociodemographic variables (race, pubertal timing or tempo, and financial strain), age, or the number of fMRI scans (Table 1). Follow-up analyses compared depression occurring prior to age 13 versus age 13 and older, as well as the effects of current total KSADS depression score (Supplemental Methods).

**Table 1:** Group comparisons of demographic characteristics and study procedures.

| | No Depression (n=125) | | Depression (n=58) | | $t/\chi^2$ | $p$ |
| --- | --- | --- | --- | --- | --- | --- |
|  | mean/N | SD/percent | mean/N | SD/percent |  |  |
| Age in years † | 18.5 | 1.35 | 18.4 | 1.4 | -0.77 | 0.44 |
| African American * | n=89 | 71% | n=44 | 76% | 0.23 | 0.63 |
| Financial strain * | n=53 | 42% | n=30 | 52% | 1.04 | 0.31 |
| Pubertal tempo x | 0.34 | 0.05 | 0.34 | 0.05 | 0.27 | 0.79 |
| Pubertal timing x | 10.25 | 0.88 | 10.20 | 0.89 | 0.47 | 0.64 |
| Number of scans x | 2.7 | 1.1 | 2.4 | 1.1 | 1.60 | 0.11 |
| Number of outlier frames † | 23.8 | 19.5 | 22.2 | 19.0 | -0.62 | 0.54 |

Descriptives and comparisons of demographic variables between participants with major depression disorder (MDD) and those without. †= As the majority of participants had data at multiple time points, group comparison was run as a linear mixed effect model controlling for participant ID as a random intercept. \*= group comparison was run as a chi-squared test. x= group comparison was run as a t-test.

### Covariates

Pubertal tempo and timing were derived from the self-reported Petersen Physical Development Scale (PPDS)^43^, administered annually from ages 9–15 years. Summary scores of girls’ pubertal maturation each year were calculated by averaging the five items. Pubertal timing (age of puberty onset) and tempo (rate of pubertal development) were estimated with a non-linear Gompertz growth model^44^.

Whether or not the participant’s household received public assistance (i.e., Women, Infants, and Children [WIC], food stamps, or welfare) during the two study visits prior to the first MRI session (i.e., ages 14 and 15) was used to determine financial strain (binary) and used as a covariate in all analyses.

### Monetary Reward and Loss fMRI Task

Participants completed a card-guessing task involving anticipation and receipt of monetary reward (Supplemental Methods, Supplemental Figure 2)^11^. Analyses focused on neural response to the cue indicating possible monetary gain (i.e., reward anticipation), given meta-analytic evidence that the correlation of MDD with striatal activation to reward anticipation, but not reward receipt, may vary with age^2^ and that striatal reward anticipation activation is predictive of future changes in depressive symptoms^13^. Details on neuroimaging data collection and processing are given in the Supplemental Methods. First-level general linear models were used to calculate the anticipation > baseline contrast. Subject-level average percent-signal-change was parcellated using the Schaefer et al. cortical atlas (n=400 regions)^45^ and the Harvard-Oxford subcortical atlas^46^ (n=24 regions). The Schaefer-atlas assigns cortical regions to one of the seven canonical resting state networks^47^. As in prior work^48^, the Neurosynth meta-analysis for ‘reward’^49^ was used to identify regions where reward-related activity is most likely to be reported (Supplemental Methods). This reward network consisted of 22 regions, including the bilateral nucleus accumbens, caudate, putamen, pallidum, amygdala, and thalamus, as well as multiple regions in the bilateral ventromedial prefrontal cortex and orbitofrontal cortex (Figure 1D). The hippocampus, the only subcortical ROI not in the reward network, was assigned to the default mode network^50^. The number of outlier frames identified during preprocessing, a metric reflecting scan quality, was used as a covariate in all analyses.

**Figure 1:**
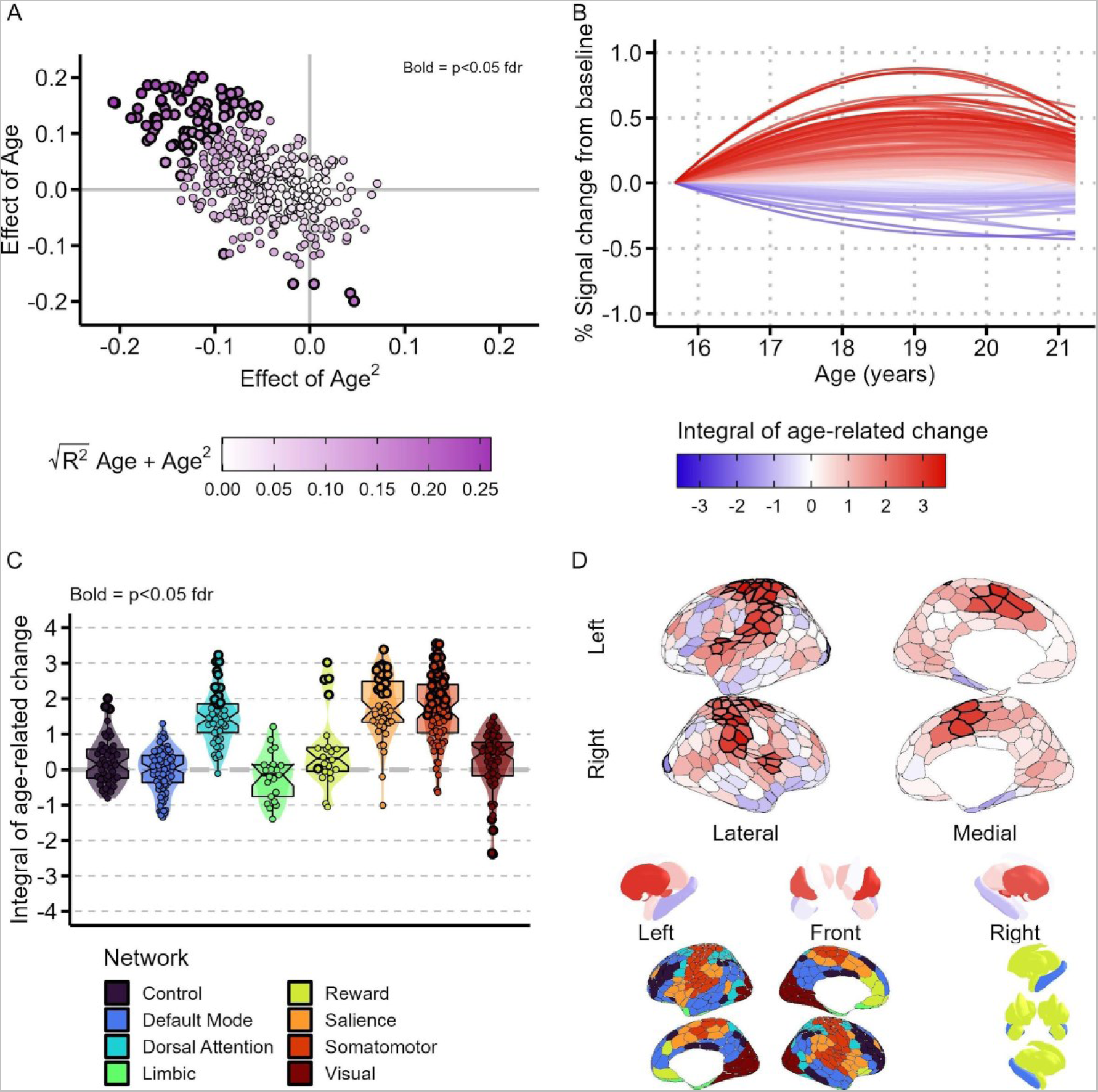
Neurodevelopment of reward anticipation-related activation in adolescent girls. **A)** Distribution of the quadratic and linear standardized effects of age on reward anticipation activation. Each point represents a different region. Significant (p<0.05 fdr-corrected) regions are bolded. The negative correlation (r=-0.59) indicates that regions with a large linear effect of age also tend to have a large negative quadratic effect (i.e., activation increases with an inverted-U trajectory, leading to a correlation as the age range is relatively narrow). Regions are colored by the square-root of the marginal R^2^ of the linear and quadratic age effects. **B**) Age-related change across all regions. Each line is a separate region, colored by the integral of the age-related change. Activation is centered so that each region begins a 0 at age 16, highlighting how activation changes with age. Most regions showed curvilinear age-related change across late adolescence, with the majority showing a positive quadratic pattern. **C**) The integral of age-related change plotted by network – each point represents a separate region. Points representing significant (p<0.05 fdr) regions are bolded. **D**) The integral of age-related change plotted by Schaefer-atlas cortical (n=400) and subcortical region. Significant (p<0.05 fdr) cortical regions have a thicker outline. Significant subcortical regions included bilateral putamen and pallidum, and the right nucleus accumbens. The figure below describes regional assignment to each network.

### Statistical analyses

#### Age

Linear mixed-effect models using maximum likelihood estimation^51, 52^ tested effects of age. Model covariates included race, pubertal tempo and timing, and financial strain. Participant ID was included as a random intercept. Age, included as a fixed effect, was modeled as a second-degree orthogonal polynomial (i.e., age + age^2^; mean-centered). Age was not modeled as a random slope, as random-slope models were singular. To determine statistical significance, a likelihood-ratio test compared a model with only covariates to a model that additionally included age and age^2^, reflecting whether the addition of age significantly improved model fit. Additional control analyses are reported in the Supplement. The effect of age on region was summarized as the integral of age-related change (Supplemental Methods).

Depression-by-Age interaction: Linear mixed-effect models tested whether the addition of depression-by-age and depression-by-age^2^ interaction terms improved model fit, using a likelihood-ratio test. The baseline model included all previously described covariates--race, pubertal tempo, and financial strain--as well as age, age^2^, and depression group as main effects. As the goal was to examine depression group differences in the correlation of activation with age, the model comparison did not test depression group differences in average activation. Models additionally included terms for the interactions between potential confounding variables and main effects (e.g., age x puberty, age x FD, etc.)^53^. Exploratory *post-hoc* analyses examined the main effect of depression, effects of depression occurring prior to age 13 versus age 13 and older, as well as effects and interactions with current depression score (Supplemental Methods).

Multiple test correction using fdr correction (Supplemental Figure 5) was jointly applied across the age and age x depression analysis results. *Post-hoc* analyses additionally compared effects between the reward and loss anticipation contrasts (Supplemental Methods).

Sensitivity analyses (Supplemental Figure 4, Supplemental Results) indicated that analyses had 80% power to detect effects as small as β = 0.1 – 0.15, with slightly less power for the MDD-by-Age interaction, likely due to imbalanced group-sizes.

## Results

### Neurodevelopment of reward anticipation activation

The addition of age and age^2^ to mixed-effect models significantly improved model-fit in n=80 regions (19.3% of 414 regions; p<0.05 fdr; ΔAIC = -30.4 --5.9, ΔBIC = -22.2 - 2.3; Figure 1A & 1B). These included regions in the somatomotor (n=37/77, 49% of regions in the network), salience (n=18/47, 38%), dorsal attention (n=13/46, 29%), reward (n=5/22, 23%), visual (n=4/61, 7%), and control (n=3/50, 6%) networks (Supplemental Data). Significant regions included the bilateral frontal operculum, insula, dorsal anterior cingulate cortex, the left parietal operculum, the bilateral frontal eye fields, the bilateral putamen (Supplemental Figure 6) and pallidum, and the right nucleus accumbens (Figure 1D). The majority of significant regions increased in activation throughout the course of the study period (Figure 1C), with the exception of regions in the visual cortex and the right nucleus accumbens (NAc), which showed a net decrease in activation, with an inverted-U shape trajectory (i.e., NAc activation peaked earlier than other significant regions). *Post-hoc* analyses indicated that these effects were not attributable to habituation^54^ (Supplemental Results). Analyses compared the derived age-associated integral of activation between networks (Supplemental Methods; Figure 2). Network-membership was significantly associated with the age-integral (F_(7,401)_=53.33, p_permutation_=4x10^-4^, adjusted R^2^=0.48). The dorsal attention, salience, and somatomotor networks showed the largest average age-integral (Figure 1C; Supplemental Table 2). Results did not differ when race was included as a covariate (Supplemental Data). *Post-hoc* analyses comparing win and loss anticipation found that age effects do not differ between these conditions (Supplemental Results).

**Figure 2.**
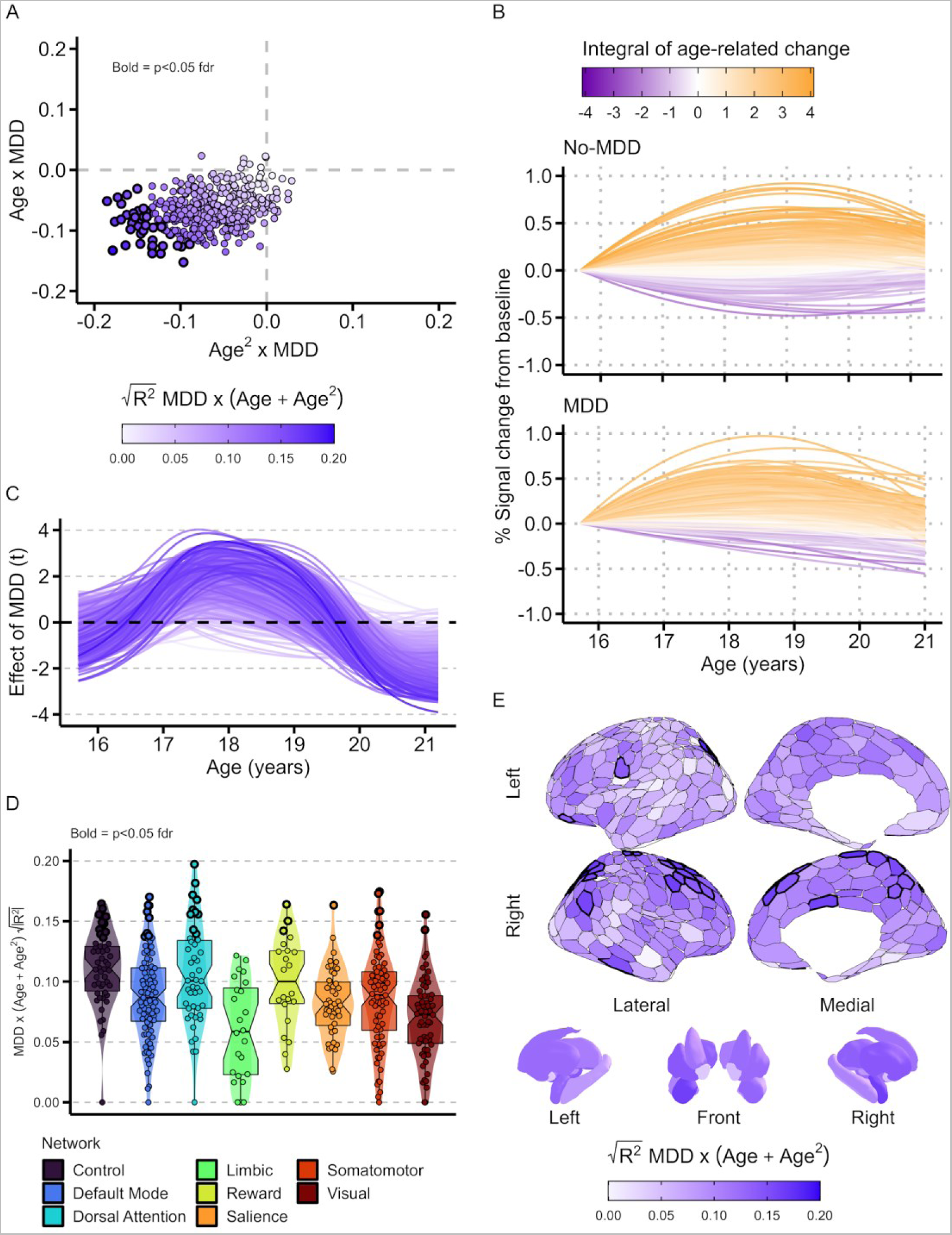
Accelerated neurodevelopment in adolescents with MDD. **A)** Distribution of the interaction effects between age (age+age^2^) and MDD (MDD n=58; No-MDD n=125). Participants with MDD display age-related change that is more negative and more concave (the correlation between linear and quadratic effects is r=0.32). Values are the standardized effect size of the interaction. Each point represents a separate region. Points are colored by the square-root of the increase in marginal R^2^ from adding the Age x MDD and Age^2^ x MDD interaction terms to the model, which is linearly proportional to the χ^2^ of the model comparison. Bolded points are significant p<0.05 fdr-corrected. **B**) Age-related changes in reward anticipation activation in participants without MDD (upper graph) and participants with MDD (lower graph). Each line represents a separate region. Line color represents the integral of the age-related change of each region. **C)**. The effect of MDD changes with age. Each line represents a different region. The y-axis is the t-statistic of the main-effect of MDD (MDD > No-MDD) when age is centered to the x-axis value. Lines are colored by the square-root of the increase in marginal R^2^ from adding the Age x MDD and Age^2^ x MDD interaction terms to the model. **D)** The square-root of the marginal R^2^ from age x MDD and age2 x MDD interactions predicting activation during reward anticipation, plotted by network. Significant (p<0.05 fdr) regions are bolded. There is no significant effect of network (p_permutation_>0.05). **E)** The square-root of the marginal R^2^ from age x MDD and age^2^ x MDD interactions predicting activation during reward anticipation, plotted by region. Significant (p<0.05 fdr) cortical regions have a thicker Outline. Significant subcortical regions included the right putamen and right amygdala.

### Neurodevelopmental trajectories and adolescent depression

The addition of interactions between MDD and age+age^2^ to mixed-effect models significantly improved model-fit in n=40 regions (9.6% of 414 regions; p<0.05 fdr; ΔAIC = -14.7 --2.6, ΔBIC = -6.5 – 5.5; Figure 2A). MDD moderated the effect of age such that MDD was associated with a more negative age-trajectory (decreased more) and a larger inverted-U shape, relative to participants with no history of MDD (Figure 2A & 2B). 91% of regions showed effects in the same direction (Figure 2A). Equivalently, the main effect of adolescent MDD changed as a function of participant age, resulting in periods of both global hyper-activation and global hypo-activation (Figure 2C). Significant regions were present in every network except the limbic network, including the control (n=10/50 regions, 20% of all regions in the network), dorsal attention (n=10/46, 22%), default mode (n=8/87, 9%), reward (n=3/22, 13.6%), salience (n=1/47, 2%), somatomotor (n=6/77, 7.8%), and visual (n=2/61, 3%) networks (Figure 2D; Supplemental Data). Regions included the right orbitofrontal cortex, right medial prefrontal cortex, right anterior cingulate, and right lateral prefrontal cortex, as well as the right amygdala and right putamen (Figure 2E; Supplemental Figure 6; Supplemental Data). Network-membership was not associated with the strength of the MDD-by-age interactions (F_(7,401)_=9.4, p_permutation_=0.42). Conclusions did not differ when race was included as a covariate (Supplemental Data).

*Post-hoc* analyses testing the main effect of MDD found a nominally significant effect in 10 regions, none of which survived fdr-correction (no region in the basal ganglia was nominally significant; Supplemental Data). *Post-hoc* analyses found no significant effect of current depression score over and above MDD, and that the MDD x Age effect remained after controlling for current depression (Supplemental Results). *Post-hoc* analyses examining age-of-onset (Supplemental Methods) found significant effects in n=12 regions (2.9% of 414 regions; p<0.05 fdr; Supplemental Figure 7). Results survived joint correction alongside the results of the primary analyses (i.e., age and MDD x age). Effects reflect a more negative quadratic effect of age among participants meeting criteria for depression prior to age 13 (i.e., the age-associated change had a greater inverted-U shape for this group; Supplemental Figure 7A). Effects of MDD in primary analyses (above) were not driven by either age-of-onset group (Supplemental Data). *Post-hoc* analyses comparing win and loss anticipation found that MDD x Age effects do not differ between these conditions (Supplemental Results).

## Discussion

Two primary findings emerged. First, while reward-related regions (e.g., the nucleus accumbens) showed evidence of age-related change in reward anticipation activation, late-adolescence was characterized predominantly by age-related change in the dorsal attention, salience (also called ventral attention), and somatomotor networks. Second, patterns of age-related change differed between participants with and without MDD, in a manner consistent with the hypothesis of accelerated neurodevelopment. Although regions that differed by MDD included canonical reward regions, such as the putamen and medial orbitofrontal cortex, effects were present in regions within every network, save the limbic network. Taken together, these findings suggest that MDD is associated with differential neurodevelopment of regions supporting reward processing, which includes many regions beyond canonical ‘core’ reward areas.

### Neurodevelopment of reward anticipation processing

A major strength of our analyses, relative to prior work, is that we tested the correlation with age, rather than comparing averages between age groups, resulting in greater sensitivity to developmental effects. Results replicate prior work identifying elevated reward-related activation in adolescence^20–22, 34–36^. While reward-related regions (i.e., bilateral putamen, bilateral pallidum, and right nucleus accumbens) exhibited age-related change, one striking observation is that reward-processing in late-adolescence was characterized predominantly by change of the salience, dorsal attention, and somatomotor networks. While these networks are not primarily associated with reward function^55^, they all include regions that contribute to reward processing and reward-driven behavior, and that display developmental changes in activation during reward processing (i.e., bilateral insula, dorsal anterior cingulate cortex, intraparietal sulcus, frontal eye fields, and motor cortex)^22, 31, 56, 57^.

The peaking of dorsal attention, salience, and motor regions in late adolescence parallels the expected developmental time-course of cognitive control^58, 59^. Dual-system models posit that adolescent reward-sensitivity is attributable to cognitive control developing later than the reward system^60^. Further, results may reflect the development of value-enhanced selective attention, which peaks in young adulthood and is thought to scaffold cognitive control^61^. That is, older adolescents may be better able to marshal attentional resources in response to a rewarding cue, as their cognitive control system is more mature. Relatedly, the present analyses focused on the reward anticipation condition, given prior evidence that striatal reward anticipation activation is predictive of future changes in depressive symptoms^13^. However, *post-hoc* analyses found that observed effects of age, and the interaction between age and MDD, did not significantly differ between reward and loss anticipation contrasts. This is broadly consistent with prior meta-analytic evidence of little or no difference between the main effects of reward and loss anticipation^62–64^. Thus, the present results may instead reflect development of the neural response to outcome magnitude, or of development of cognitive control or attention to salient events, rather than development of the response to cue valence (i.e., reward vs. loss). However, as our task did not include cognitive control or attentional demands, or rewards of differing magnitudes, we could not test this hypothesis.

### Accelerated reward anticipation neurodevelopment in adolescents with depression

Participant age interacted with MDD history in n=40 regions, such that participants with MDD had group-level developmental trajectories that were more negative and more concave (e.g., a greater ‘inverted-U’ shape), but did not differ in average activation (Figure 2). Strikingly, 91% of all regions showed a similar pattern of negative age effects both linearly and quadratically (Figure 2A), and comparison between networks suggest that these effects are not specific to any network. These results suggest that MDD is associated with accelerated whole-brain neurodevelopment of reward anticipation-related processes in late adolescence.

Regions with a significant interaction between MDD and age included three with well-established roles in reward processing: the right putamen (Supplemental Figure 6), a sub-region of the right medial orbitofrontal cortex (OFC), and the right amygdala, (Supplemental Data)^3, 65^. This finding converges with prior meta-analytic evidence for blunted putamen activation during reward anticipation in participants with current MDD^2^, including the observation that it is associated with cumulative depression severity in a sample of younger adolescents oversampled for preschool-onset depression^66^. Accelerated structural aging of the putamen has additionally been reported in a sample of adults with MDD^67^. There is also meta-analytic evidence supporting increased medial OFC activation during reward processing in MDD, in a cluster that is proximal to the observed significant ROI^3^.

Significant interactions between age and MDD were also present in every network, save the limbic. While networks did not differ in their mean effect, the majority of significant regions were located either in the control (n=15) or dorsal attention (n=12) networks. All but one of the regions in the dorsal attention network were located in the posterior parietal cortex, which plays a role in reward-driven attentional control^68^. Regions in the control network included sub-regions of the lateral prefrontal cortex, posterior cingulate, inferior parietal sulcus, precuneus, and posterior medial prefrontal cortex. These regions all play key roles in cognitive control^55^, which uses reward information to guide actions^69, 70^. Depression is known to be associated with deficits in executive function^71^, though there is limited evidence of differential activation of cognitive control regions in youth with depression^72^.

The present results support a model of accelerated system-wide neurodevelopment in reward processing in relation to depression, and point to the need for prospective studies (or for leveraging ongoing large-scale studies) that cover a larger age-range. This finding converges with growing evidence of accelerated structural aging and development in depression across the lifespan^67, 73, 74^. However, studies of childhood- or adolescent-onset depression have been mixed^75^. *Post-hoc* findings of a more curvilinear developmental trajectory of reward-related activation in adolescents with MDD that onset at ages 9-12 years compared with adolescents with MDD that onset at ages 13-20 years (Supplemental Figure 7A) indicate possible greater acceleration of brain development in girls with earlier age of MDD occurrence. This suggests that age-of-onset may be an important aspect of depression to consider when examining adolescent neurodevelopment of reward processing specifically in girls. More broadly, this result suggests that accelerated neurodevelopment of reward processing may be in response to factors which increase risk for depression early in life, such as adversity^76, 77^. For example, accelerated neurodevelopment of reward may lead to an earlier increase in sensation seeking during adolescence^78^, which is hypothesized to promote adaptive responses to stress^79^.

### Reward disruption emerges from differential neurodevelopment

The present results challenge the perspective that depression emerges from stable disruption of reward network function, particularly blunted VS activation and hyperactivation of the OFC^1, 3, 13^. We observed both nearly-global hyperactivation and nearly-global hypoactivation in participants with MDD, depending on age (Figure 2C). Disrupted reward processing seen cross-sectionally in MDD could emerge from accelerated neurodevelopment in late adolescence (Supplemental Figure 6)^5^. Notably, differential neurodevelopment is not restricted to classic reward regions. Disruptions to development may have consequences both for cognitive function and for the emergence of reward-system-specific function. For example a shortened or earlier window of elevated adolescent reward-sensitivity may alter motivation for rewards or pathways between cognitive control and reward circuitry^60^.

### Limitations

We cannot establish whether neurodevelopmental acceleration is a cause, consequence, or epiphenomenon of MDD, as MRI scanning only occurred during late adolescence. Large longitudinal studies will be needed to distinguish between these possibilities. We were also unable to identify whether developmental acceleration occurred before or during the study period. Additionally, the time frame for assessing depression varied across age (past month to past year), and for all participants there were between 2 and 7 years (median 3 years) during which assessments of depression did not occur. As this study was conducted exclusively in adolescent girls, further work will be needed to assess the generalizability to boys.

## Conclusions

In contrast to neurodevelopmental theories of the contribution of reward processing to adolescent risk for depression that focus primarily on stable disruption in core reward regions (e.g., the nucleus accumbens and OFC)^1, 21, 60^, the present results demonstrate that reward-processing in late adolescence undergoes age-related changes in activation in a host of regions including, but not limited to, core reward areas and that depression is associated with differing trajectories, consistent with the hypothesis of accelerated neurodevelopment. We propose that developmental accounts of the effects of reward function on psychopathology should consider the role of a broader set of “non-reward” regions, which may contribute to the effects of risk factors on the etiology and pathophysiology of psychopathology.

## Acknowledgements

The authors would like to thank the research participants. This work was supported by grants from the National Institutes of Mental Health, including MH093605 (Guyer, Keenan, Forbes), and DB was supported by T32 MH018951 (Goldstein).

## Data availability

Summary statistics generated by these analyses are provided in the Supplemental Data file. Formal data sharing proposal and agreement forms for the Pittsburgh Girls Study - Emotions Substudy can be requested from senior authors Dr. Keenan or Dr. Hipwell.

## References

1. Forbes EE, Dahl RE. Altered Reward Function in Adolescent Depression: What, When, and How? J Child Psychol Psychiatry. 2012;53(1):3–15.

2. Keren H, O’Callaghan G, Vidal-Ribas P, et al. Reward Processing in Depression: A Conceptual and Meta-Analytic Review Across fMRI and EEG Studies. Am J Psychiatry. 2018;175(11):1111–1120.

3. Ng TH, Alloy LB, Smith DV. Meta-analysis of reward processing in major depressive disorder reveals distinct abnormalities within the reward circuit. Transl Psychiatry. 2019;9(1):293.

4. Telzer EH, Fuligni AJ, Lieberman MD, Galván A. Neural sensitivity to eudaimonic and hedonic rewards differentially predict adolescent depressive symptoms over time. Proc Natl Acad Sci U S A. 2014;111(18):6600–6605.

5. Hanson JL, Hariri AR, Williamson DE. Blunted ventral striatum development in adolescence reflects emotional neglect and predicts depressive symptoms. Biol Psychiatry. 2015;78(9):598–605.

6. Stringaris A, Vidal-Ribas Belil P, Artiges E, et al. The Brain’s Response to Reward Anticipation and Depression in Adolescence: Dimensionality, Specificity, and Longitudinal Predictions in a Community-Based Sample. Am J Psychiatry. 2015;172(12):1215–1223.

7. Gotlib IH, Hamilton JP, Cooney RE, Singh MK, Henry ML, Joormann J. Neural processing of reward and loss in girls at risk for major depression. Arch Gen Psychiatry. 2010;67(4):380–387.

8. Luking KR, Pagliaccio D, Luby JL, Barch DM. Depression Risk Predicts Blunted Neural Responses to Gains and Enhanced Responses to Losses in Healthy Children. J Am Acad Child Adolesc Psychiatry. 2016;55(4):328–337.

9. Olino TM, McMakin DL, Morgan JK, et al. Reduced reward anticipation in youth at high-risk for unipolar depression: A preliminary study. Dev Cogn Neurosci. 2014;8:55–64.

10. Sharp C, Kim S, Herman L, Pane H, Reuter T, Strathearn L. Major depression in mothers predicts reduced ventral striatum activation in adolescent female offspring with and without depression. J Abnorm Psychol. 2014;123(2):298–309.

11. Forbes EE, Olino TM, Ryan ND, et al. Reward-Related Brain Function as a Predictor of Treatment Response in Adolescents with Major Depressive Disorder. Cogn Affect Behav Neurosci. 2010;10(1):107–118.

12. Queirazza F, Fouragnan E, Steele JD, Cavanagh J, Philiastides MG. Neural correlates of weighted reward prediction error during reinforcement learning classify response to cognitive behavioral therapy in depression. Sci Adv. 2019;5(7):eaav4962.

13. Nielson DM, Keren H, O’Callaghan G, et al. Great Expectations: A Critical Review of and Suggestions for the Study of Reward Processing as a Cause and Predictor of Depression. Biol Psychiatry. 2020;0(0).

14. Alexander L, Gaskin PLR, Sawiak SJ, et al. Fractionating Blunted Reward Processing Characteristic of Anhedonia by Over-Activating Primate Subgenual Anterior Cingulate Cortex. Neuron. 2019;101(2):307–320.e6.

15. Nestler EJ, Carlezon WA. The Mesolimbic Dopamine Reward Circuit in Depression. Biol Psychiatry. 2006;59(12):1151–1159.

16. Kwong ASF, Morris TT, Timpson NJ, et al. Polygenic Risk for Depression is Associated with the Severity and Rate of Change in Depressive Symptoms Across Adolescence. medRxiv. Published online May 4, 2020:2019.12.31.19016212.

17. Rohde P, Lewinsohn PM, Klein DN, Seeley JR, Gau JM. Key Characteristics of Major Depressive Disorder Occurring in Childhood, Adolescence, Emerging Adulthood, and Adulthood: Clin Psychol Sci. Published online October 17, 2012.

18. Thapar A, Collishaw S, Pine DS, Thapar AK. Depression in adolescence. The Lancet. 2012;379(9820):1056–1067.

19. Breslau J, Gilman SE, Stein BD, Ruder T, Gmelin T, Miller E. Sex differences in recent first-onset depression in an epidemiological sample of adolescents. Transl Psychiatry. 2017;7(5):e1139–e1139.

20. Braams BR, van Duijvenvoorde ACK, Peper JS, Crone EA. Longitudinal Changes in Adolescent Risk-Taking: A Comprehensive Study of Neural Responses to Rewards, Pubertal Development, and Risk-Taking Behavior. J Neurosci. 2015;35(18):7226–7238.

21. Kujawa A, Klein DN, Pegg S, Weinberg A. Developmental trajectories to reduced activation of positive valence systems: A review of biological and environmental contributions. Dev Cogn Neurosci. 2020;43:100791.

22. Silverman MH, Jedd K, Luciana M. Neural networks involved in adolescent reward processing: An activation likelihood estimation meta-analysis of functional neuroimaging studies. NeuroImage. 2015;122:427–439.

23. Raznahan A, Shaw PW, Lerch JP, et al. Longitudinal four-dimensional mapping of subcortical anatomy in human development. Proc Natl Acad Sci. 2014;111(4):1592–1597.

24. Shulman EP, Harden KP, Chein JM, Steinberg L. Sex differences in the developmental trajectories of impulse control and sensation-seeking from early adolescence to early adulthood. J Youth Adolesc. 2015;44(1):1–17.

25. Pagliaccio D, Luking KR, Anokhin AP, et al. Revising the BIS/BAS to Study Development: Measurement Invariance and Normative Effects of Age and Sex from Childhood through Adulthood. Psychol Assess. 2016;28(4):429–442.

26. Luking KR, Pagliaccio D, Luby JL, Barch DM. Reward Processing and Risk for Depression Across Development. Trends Cogn Sci. 2016;20(6):456–468.

27. Bos MGN, Peters S, Van De Kamp FC, Crone EA, Tamnes CK. Emerging depression in adolescence coincides with accelerated frontal cortical thinning. J Child Psychol Psychiatry. 2018;59(9):994–1002.

28. Callaghan BL, Tottenham N. The Stress Acceleration Hypothesis: effects of early-life adversity on emotion circuits and behavior. Curr Opin Behav Sci. 2016;7:76–81.

29. Haber SN, Knutson B. The reward circuit: linking primate anatomy and human imaging. Neuropsychopharmacol Off Publ Am Coll Neuropsychopharmacol. 2010;35(1):4–26.

30. Schultz W. Multiple Reward Signals. Nat Rev Neurosci. 2000;1(December):199–207.

31. Cao Z, Bennett M, Orr C, et al. Mapping adolescent reward anticipation, receipt, and prediction error during the monetary incentive delay task. Hum Brain Mapp. 2019;40(1):262–283.

32. Vickery TJ, Chun MM, Lee D. Ubiquity and Specificity of Reinforcement Signals throughout the Human Brain. Neuron. 2011;72(1):166–177.

33. O’Callaghan G, Stringaris A. Reward processing in adolescent depression across neuroimaging modalities: A review. Z Für Kinder-Jugendpsychiatrie Psychother. 2019;47(6):535–541.

34. Galvan A, Hare TA, Parra CE, et al. Earlier Development of the Accumbens Relative to Orbitofrontal Cortex Might Underlie Risk-Taking Behavior in Adolescents. J Neurosci. 2006;26(25):6885–6892.

35. McCormick EM, Telzer EH. Adaptive Adolescent Flexibility: Neurodevelopment of Decision-making and Learning in a Risky Context. J Cogn Neurosci. 2016;29(3):413–423.

36. Van Leijenhorst L, Moor BG, Op de Macks ZA, Rombouts SARB, Westenberg PM, Crone EA. Adolescent risky decision-making: Neurocognitive development of reward and control regions. NeuroImage. 2010;51(1):345–355.

37. Ridley M, Rao G, Schilbach F, Patel V. Poverty, depression, and anxiety: Causal evidence and mechanisms. Science. 2020;370(6522):eaay0214.

38. Daly M. Prevalence of Depression Among Adolescents in the U.S. From 2009 to 2019: Analysis of Trends by Sex, Race/Ethnicity, and Income. J Adolesc Health. 2022;70(3):496–499.

39. Verhoeven JE, Révész D, Epel ES, Lin J, Wolkowitz OM, Penninx BWJH. Major depressive disorder and accelerated cellular aging: results from a large psychiatric cohort study. Mol Psychiatry. 2014;19(8):895–901.

40. Keenan K, Hipwell A, Chung T, et al. The Pittsburgh Girls Study: overview and initial findings. J Clin Child Adolesc Psychol Off J Soc Clin Child Adolesc Psychol Am Psychol Assoc Div 53. 2010;39(4):506–521.

41. Keenan K, Hipwell A, Feng X, et al. Subthreshold symptoms of depression in preadolescent girls are stable and predictive of depressive disorders. J Am Acad Child Adolesc Psychiatry. 2008;47(12):1433–1442.

42. Kaufman J, Birmaher B, Brent D, et al. Schedule for Affective Disorders and Schizophrenia for School-Age Children-Present and Lifetime Version (K-SADS-PL): initial reliability and validity data. J Am Acad Child Adolesc Psychiatry. 1997;36(7):980–988.

43. Petersen AC, Crockett L, Richards M, Boxer A. A self-report measure of pubertal status: Reliability, validity, and initial norms. J Youth Adolesc. 1988;17(2):117–133.

44. Chahal R, Vilgis V, Grimm KJ, et al. Girls’ pubertal development is associated with whitematter microstructure in late adolescence. NeuroImage. 2018;181:659–669.

45. Schaefer A, Kong R, Gordon EM, et al. Local-Global Parcellation of the Human Cerebral Cortex from Intrinsic Functional Connectivity MRI. Cereb Cortex. Published online 2017:1–20.

46. Frazier JA, Chiu S, Breeze JL, et al. Structural brain magnetic resonance imaging of limbic and thalamic volumes in pediatric bipolar disorder. Am J Psychiatry. 2005;162(7):1256–1265.

47. Yeo BTT, Krienen FM, Sepulcre J, et al. The organization of the human cerebral cortex estimated by intrinsic functional connectivity. J Neurophysiol. 2011;106(3):1125–1165.

48. Baranger DAA, Lindenmuth M, Nance M, et al. The longitudinal stability of fMRI activation during reward processing in adolescents and young adults. NeuroImage. 2021;232:117872.

49. Yarkoni T, Poldrack RA, Nichols TE, Van Essen DC, Wager TD. Large-scale automated synthesis of human functional neuroimaging data. Nat Methods. 2011;8(8):665–670.

50. Buckner RL, Andrews-Hanna JR, Schacter DL. The brain’s default network: anatomy, function, and relevance to disease. Ann N Y Acad Sci. 2008;1124:1–38.

51. Bates D, Mächler M, Bolker BM, Walker SC, Maechler Martin, Walker SC. Fitting linear mixed-effects models using lme4. J Stat Softw. 2015;67(1):1–48.

52. R Core Team. R: A Language and Environment for Statistical Computing. Published online 2021.

53. Keller MC. Gene × environment interaction studies have not properly controlled for potential confounders: the problem and the (simple) solution. Biol Psychiatry. 2014;75(1):18–24.

54. Hasler BP, Forbes EE, Franzen PL. Time-of-day differences and short-term stability of the neural response to monetary reward: A pilot study. Psychiatry Res Neuroimaging. 2014;224(1):22–27.

55. Dosenbach NUF, Fair DA, Miezin FM, et al. Distinct brain networks for adaptive and stable task control in humans. Proc Natl Acad Sci. 2007;104(26):11073–11078.

56. Hoogendam JM, Kahn RS, Hillegers MHJ, van Buuren M, Vink M. Different developmental trajectories for anticipation and receipt of reward during adolescence. Dev Cogn Neurosci. 2013;6:113–124.

57. Yaple ZA, Yu R, Arsalidou M. Spatial migration of human reward processing with functional development: Evidence from quantitative meta-analyses. Hum Brain Mapp. 2020;41(14):3993–4009.

58. Kim-Spoon J, Herd T, Brieant A, et al. A 4-year longitudinal neuroimaging study of cognitive control using latent growth modeling: developmental changes and brain-behavior associations. NeuroImage. 2021;237:118134.

59. Luna B. DEVELOPMENTAL CHANGES IN COGNITIVE CONTROL THROUGH ADOLESCENCE. Adv Child Dev Behav. 2009;37:233–278.

60. Shulman EP, Smith AR, Silva K, et al. The dual systems model: Review, reappraisal, and reaffirmation. Dev Cogn Neurosci. 2016;17:103–117.

61. Davidow JY, Insel C, Somerville LH. Adolescent Development of Value-Guided Goal Pursuit. Trends Cogn Sci. 2018;22(8):725–736.

62. Chen Y, Chaudhary S, Li CSR. Shared and distinct neural activity during anticipation and outcome of win and loss: A meta-analysis of the monetary incentive delay task. NeuroImage. 2022;264:119764.

63. Wilson RP, Colizzi M, Bossong MG, et al. The Neural Substrate of Reward Anticipation in Health: A Meta-Analysis of fMRI Findings in the Monetary Incentive Delay Task. Neuropsychol Rev. 2018;28(4):496–506.

64. Oldham S, Murawski C, Fornito A, Youssef G, Yücel M, Lorenzetti V. The anticipation and outcome phases of reward and loss processing: A neuroimaging meta-analysis of the monetary incentive delay task. Hum Brain Mapp. 2018;39(8):3398–3418.

65. Long Y, Cao H, Yan C, et al. Altered resting-state dynamic functional brain networks in major depressive disorder: Findings from the REST-meta-MDD consortium. NeuroImage Clin. 2020;26:102163.

66. Rappaport BI, Kandala S, Luby JL, Barch DM. Brain Reward System Dysfunction in Adolescence: Current, Cumulative, and Developmental Periods of Depression. Am J Psychiatry. 2020;177(8):754–763.

67. Sacchet MD, Camacho MC, Livermore EE, Thomas EAC, Gotlib IH. Accelerated aging of the putamen in patients with major depressive disorder. J Psychiatry Neurosci JPN. 2017;42(3):164–171.

68. Peck CJ, Jangraw DC, Suzuki M, Efem R, Gottlieb J. Reward modulates attention independently of action value in posterior parietal cortex. J Neurosci Off J Soc Neurosci. 2009;29(36):11182–11191.

69. Duverne S, Koechlin E. Rewards and Cognitive Control in the Human Prefrontal Cortex. Cereb Cortex. 2017;27(10):5024–5039.

70. Soltani A, Koechlin E. Computational models of adaptive behavior and prefrontal cortex. Neuropsychopharmacology. 2022;47(1):58–71.

71. Snyder HR. Major depressive disorder is associated with broad impairments on neuropsychological measures of executive function: A meta-analysis and review. Psychol Bull. 2013;139(1):81–132.

72. Miller CH, Hamilton JP, Sacchet MD, Gotlib IH. Meta-analysis of Functional Neuroimaging of Major Depressive Disorder in Youth. JAMA Psychiatry. 2015;72(10):1045–1053.

73. Ballester PL, Romano MT, de Azevedo Cardoso T, et al. Brain age in mood and psychotic disorders: a systematic review and meta-analysis. Acta Psychiatr Scand. 2022;145(1):42–55.

74. Han LKM, Dinga R, Hahn T, et al. Brain aging in major depressive disorder: results from the ENIGMA major depressive disorder working group. Mol Psychiatry. 2021;26(9):5124–5139.

75. Nielsen JD, Mennies RJ, Olino TM. Application of a diathesis-stress model to the interplay of cortical structural development and emerging depression in youth. Clin Psychol Rev. 2020;82:101922.

76. Kessler RC, Davis CG, Kendler KS. Childhood adversity and adult psychiatric disorder in the US National Comorbidity Survey. Psychol Med. 1997;27(5):1101–1119.

77. Romens SE, Casement MD, McAloon R, et al. Adolescent girls’ neural response to reward mediates the relation between childhood financial disadvantage and depression. J Child Psychol Psychiatry. 2015;56(11):1177–1184.

78. Chase HW, Fournier JC, Bertocci MA, et al. A pathway linking reward circuitry, impulsive sensation-seeking and risky decision-making in young adults: identifying neural markers for new interventions. Transl Psychiatry. 2017;7(4):e1096.

79. Norbury A, Husain M. Sensation-seeking: Dopaminergic modulation and risk for psychopathology. Behav Brain Res. 2015;288:79–93.

