## Supplement for "Accelerated neurodevelopment of reward anticipation processing in adolescent girls with depression"

**Table of Contents:**

Supplemental Methods: pg. 3-6

Supplemental Results: pg. 6-8

Supplemental Tables: pg. 9

Supplemental Figures: pgs. 10-16

**Supplemental Methods**

*Early-onset Depression*: Participants with lifetime depression (N=58) were split into two groups – those with an earlier age of onset (at or before the age of 12; n= 34) and those with a later age of onset (on or after the fourth wave of data collection, age 13; n=24), up to age 20. This cut-point was chosen as it was when participants had on average reached Tanner stage 3[1], which is considered a threshold for pubertal maturation. Depression age of onset was also binarized, as for many participants (46%) the precise age of onset is unknown, owing either to depression onset occurring prior to the first wave of data collection at age 9 (n=25), or missing data (i.e., the first wave when the participant met criteria was after a wave with missing data, n=2). Participants with earlier-onset MDD did not differ from those with later-onset MDD in any demographic variable, age, or the number of fMRI scanning sessions (Supplemental Table 1). *Post-hoc* age x depression-onset analysis results were jointly corrected with the results of the primary age and age x depression results (n=1,242 tests in total).

*Monetary Reward and Loss fMRI Task:* Participants completed an 8-minute slow event-related card-guessing task involving anticipation and receipt of monetary reward (Supplemental Figure 2) [2]. This task occurred approximately 12 minutes into a 50 minute scanning session. In each trial (20 s) participants were shown a card with a possible value of 1–9, and asked to guess whether it was lower or higher than 5 (4 s). Participants then learned the trial-type (6 s) – possible-win or possible-loss (i.e., the anticipation phase). Participants were told the "correct" answer (500 msec) and then whether they had a positive (gain money), negative (lose money), or neutral (no-change) outcome (500 msec; i.e., the outcome phase). Each trial ended with a cross-hair that was presented during a 9 second inter-trial interval. Participants completed 24 trials, with a balanced number of trial types (i.e., 12 possible-win and 12 possible-loss). Trials were presented in a pseudorandom order with predetermined outcomes. Participants received $10 after completing the task.

*Neuroimaging data collection and preprocessing*
Data collection: Participants were scanned using a Siemens 3T Trio scanner. BOLD functional images were acquired with a gradient echo planar imaging (EPI) sequence and covered 39 axial slices (3.1 mm wide; no gap) beginning at the cerebral vertex and extending across the entire cerebrum and the majority of the cerebellum (TR/RE = 2000/25ms, field of view = 20cm, matrix = 64×64). A reference EPI scan was acquired before fMRI data collection, which was visually inspected for artifacts (e.g., ghosting) and for adequate signal across the entire volume. A 160-slice high-resolution sagittally acquired T1-weighted anatomical image (MPRAGE) was collected for co-registration and normalization of functional images (TR/TE = 2300/2.98 ms, flip angle=90°, field of view = 20 cm, matrix = 256 × 240). No acceleration was used.

Preprocessing and within-person analysis: Preprocessing was completed using Statistical Parametric Mapping software (SPM8; http://www.fil.ion.ucl.ac.uk/spm). Structural images for each participant were auto-segmented, slice timing corrected to the middle volume of the time-series, and functional images were realigned to correct for head motion, registered to the segmented structural data, spatially normalized into standard stereotaxic space (Montreal Neurological Institute template) using a 12-parameter affine model, and smoothed with a 6mm full-width at half-maximum Gaussian filter. Voxel-wise signal was ratio-normalized to the whole-brain global mean. The Artifact Detection Toolbox (ART; http://www.nitrc.org/projects/artifact_detect/) software was used to detect functional volumes with movement > 3 SD from the subject's mean, > 0.5 mm scan-to-scan translation, or > 0.01 degrees of scan-to-scan rotation. Preprocessed data were inspected to ensure that all participants had fewer than 25% of volumes with excessive movement detected by ART, good scan quality, and ventral striatum coverage of at least 80%. Temporal censoring based on ART output was used to remove motion artifacts in first-level analyses. First-level general linear models (GLMs) were used to calculate images for the in anticipation > baseline contrast. Reward anticipation was defined as the 6-seconds when the symbol indicating trial-type was displayed, the 1 second of outcome, and the first second of fixation (8 seconds total), to account for the delay in hemodynamic response relative to neural activity and capture as much of the reward anticipation response as possible while avoiding substantial overlap with BOLD response to reward outcome events. Baseline was defined as the last 3 seconds of the inter-trial interval.

Cortical regions of interest (ROIs) were defined using the Schaefer atlas[3], a recent cortical parcellation derived from resting-state fMRI data, which also assigns each ROI to one of seven canonical resting state networks[4]. The 400-region version of the Schaefer atlas was used, as it has been estimated that there are approximately 300-400 cortical regions[5]. Subcortical ROIs were identified using the Harvard-Oxford subcortical atlas[6] thresholded to 25% probability, and voxels present across multiple ROIs were discarded. Subject-level average percent-signal-change was extracted from each ROI, using the Marsbar toolbox for SPM[7]. The 0%, 25%, 50%, 75%, and 100% quantiles for the correlation between regions are [0.13 0.30 0.39 0.48 0.96]. As in prior work[8], the Neurosynth meta-analysis for 'reward'[9,10] was used to identify ROIs where reward-related activity is most likely to be reported. This reward network consisted of 22 regions, with bilateral subcortical regions including the bilateral nucleus accumbens, caudate, putamen, pallidum, amygdala, and thalamus. Cortical reward-network regions included multiple ROIs in the bilateral ventromedial prefrontal cortex and orbitofrontal cortex. All other cortical ROIs retained their original assignment to one of the seven resting state networks and the hippocampus, the only subcortical ROI not assigned to the reward network, was manually assigned to the limbic network, as it has long been recognized as a central component of that system[11].

As in prior work[8], an eighth 'reward' network was defined using Neurosynth[10]. Neurosynth is a platform which generates meta-analyses, using reported loci from published fMRI studies, thus identifying the regions of the brain most likely to be reported in studies on a given topic. We used the Neurosynth meta-analysis of reward-associated keywords, including "reward", "outcome", "anticipation", and "monetary" (Supplemental Figure 3; e.g. topic 7 from the v5 50-topic solution[9], which identifies voxels ($p<0.01$ false-discovery-rate corrected) more likely to be reported in reward studies (n=1,218) than non-reward studies (n=13,153). https://neurosynth.org/analyses/topics/v5-topics-50/7). ROIs were assigned to the reward network based on the overlap between voxels in the Neurosynth meta-analysis and each ROI. A t-statistic was calculated based on the distribution of this overlap, and regions that showed a significant overlap at $p<0.05$ were assigned to the reward network.

Sensitivity analyses: Simulation-based sensitivity analyses were conducted using the simr R-package[12]. Effects of interest (Age and $Age^2$, or MDD x Age + $Age^2$) were iteratively increased from 0 to 0.2 in increments of 0.01. 5,000 simulations were run per setting combination. Power was determined as the percent of simulations where the log-likelihood ratio test, comparing a model with the effects of interest to a model without the effects of interest (as was done in primary analyses), was significant.

*Summary measure of development*: Developmental trajectories for each region were summarized using the integral of the quadratic polynomial described by fixed-effects. For this measure, activation of each region was re-centered so that the model intercept at the group-level minimum age was zero. The linear mixed-effect model was then refit, and the integral was calculated from the second-order polynomial described by the linear and quadratic fixed-effects of age. This integral represents the net-total group-level activation change over the course of the study.

Habituation: *post-hoc* analyses were conducted to test for alternative explanations for the observed main-effects of age in primary analyses. These include non-developmental effects, such as response habituation to multiple presentations of the same fMRI task and regression to the mean. Two control analyses were conducted to assess whether these effects may be present. First, analyses testing the main-effect of age were repeated, using only the first fMRI scan from each participant. Linear models were used in place of mixed-effect models. Statistical significance was not assessed (owing to the substantive reduction in number of observations), instead effect sizes were compared across the two sets of analyses. A high correlation between the estimated effects would indicate that results are not primarily attributable to non-developmental effects. Second, to assess whether habituation effects are present, visit-number and visit-number$^2$ were added to mixed effect models which already included age and age$^2$. A likelihood ratio test was used to evaluate whether the addition of these variables significantly improved model fit, with q-value fdr correction for multiple comparisons. Visit-number was not included in primary analyses as it is highly collinear with participant age ($r$=0.72, $p$<2.2x10$^{-16}$); including it would dramatically reduce the precision of estimates of age-effects.

Current depression: *post-hoc* analyses tested whether there was an effect of current depression score (i.e., KSADS depression severity at a given imaging wave) over and above depression history, whether current depression score interacted with age, controlling for depression history, and conversely whether the observed effect of depression history remained after including current depression, and the interaction between current depression and Age + Age$^2$, as covariates. These analyses were conducted identically to primary analyses.

Effect of contrast: *post-hoc* analyses compared win anticipation (the contrast used in primary analyses) to loss anticipation. These analyses tested whether the effect of Age + Age$^2$ interacted with contrast (i.e., whether the effect of age differed between the two contrasts), and whether the effect of the MDD x Age + Age$^2$ interacted with contrast (a three-way interaction; whether the interaction between MDD and Age differed between the two contrasts). Analyses were run by comparing models with the terms of interest to models without, as in primary analyses.

Network associations: Follow-up analyses tested whether resting-state networks were associated with regional results (e.g., whether effects are stronger in some networks). However, due to spatial auto-correlation, regions cannot be treated as independent observations. Instead, the analysis pipeline was repeated for 5,000 permutations of age (thus maintaining the spatial auto-correlation of regions), and significance was approximated by a gamma distribution[13]. Permutations which did not violate the multi-level structure of the data (i.e., multiple observations per participant) were identified using the R package 'permute'[14,15].

Visualization of results: As primary results consist of the comparison between two models (e.g., models with and without 'age + age$^2$'), the $\chi^2$ statistic is the effect size of interest. To facilitate interpretation, figures instead use the square-root of the difference in the marginal pseudo-$R^2$ between the two models (i.e., approximately how much more variance is explained when 'age + age$^2$' is added to the model) [16], since the $R^2$ statistic is linearly proportional to the $\chi^2$ statistic.

**Supplemental Results**

Sensitivity analyses: Sensitivity analyses found that primary analyses had 80% power to detect effects as small as β = 0.1 – 0.15, and that *post-hoc* analyses comparing age-of-onset within the MDD group had had 80% power to detect effects as small as β = 0.2 – 0.27 (Supplemental Figure 4). The sensitivity analysis for the interaction between MDD and Age + Age$^2$ yielded slightly larger effect sizes than the sensitivity analysis for Age + Age$^2$, which is likely due to the imbalance in group sizes[17]. It should be emphasized that sensitivity analyses (or indeed any power analysis) cannot be used on its own to infer whether or not results are false-positives. Rather, as our observed significant effects are in many cases near (or in the case of *post-hoc* age-of-onset analyses – below) our smallest detectable effects size, we can infer that there is a high probability that many of our observed significant effects are over estimates of the true population effect - an extremely common scenario. Thus, future work which may seek to replicate or extend the present results should plan on detecting effect sizes that are smaller than those reported here.

Habituation: Control analyses first tested whether developmental effects remained when only the first fMRI scan from each participant was included. If development is not driving observed age-associated changes, then estimates from these control analyses would be expected to be uncorrelated with results which include every fMRI scan session. The fixed-effects of age, age$^2$, the model intercept, and the integral of age-associated changes, were all highly correlated with their original estimates, when only the first scan was included (paired correlation; age: $r=0.79, p<2.2\times10^{-16}$; age$^2$: $r=0.70$ $p<2.2\times10^{-16}$; intercept: $r=0.71$, $p<2.2\times10^{-16}$; integral: $r=0.99$, $p<2.2\times10^{-16}$). Second, it was tested whether the addition of fixed effects for scan-number and scan-number$^2$ (i.e., habituation effects) improved model fit. While these effects were nominally significant in 25 regions, no region survived fdr-correction for multiple comparisons (Supplemental Data). Overall, these results indicate that habituation or regression to the mean are not the primary drivers of the observed developmental effects in this study.

Current KSADS x MDD: 3 regions were nominally significant in analyses of current KSADS score (controlling for MDD), and 11 were nominally significant in analyses of the interaction between current KSADS score and age (controlling for the MDD x age interaction) – none of which survive multiple test correction (Supplemental Data). In contrast, 52 regions showed a significant interaction between depression and age after controlling for total current KSADS score and its interaction with age. These results indicate that our findings are largely independent of current depression, and may be more specific to depression history or risk for depression.

Win vs. Loss anticipation: Task condition (win vs. loss) did not interact with age (smallest uncorrected p-value = 0.23; Supplemental Data). Similarly, there was no three-way interaction between condition, depression, and age (smallest uncorrected p-value = 0.34; Supplemental Data). These results indicate 1) that age-related changes in loss anticipation do not significantly differ from our current results, and 2) that the difference between win and loss anticipation (i.e., win > loss) does not vary with age. This conclusion is less surprising when we consider the high correlation between win and loss anticipation – the 0%, 25%, 50%, 75%, and 100% quantiles are 0.74, 0.84, 0.85, 0.87, and 0.93. This is not to say that the average activation does not differ between conditions – indeed the bilateral nucleus accumbens (NAc) shows a large effect of condition (win>loss), larger than any other region (right NAc: t=6.1, p=1.4x10-9; left NAc: t=5.5, p=5.6x10-8). Rather, while there is a mean difference between conditions, the rank-order of individuals across regions is highly consistent between conditions.

**References**

1. Chahal R, Vilgis V, Grimm KJ, et al. Girls' pubertal development is associated with white matter microstructure in late adolescence. *NeuroImage*. 2018;181:659-669. doi:10.1016/j.neuroimage.2018.07.050
2. Forbes EE, Olino TM, Ryan ND, et al. Reward-Related Brain Function as a Predictor of Treatment Response in Adolescents with Major Depressive Disorder. *Cogn Affect Behav Neurosci*. 2010;10(1):107-118. doi:10.3758/CABN.10.1.107
3. Schaefer A, Kong R, Gordon EM, et al. Local-Global Parcellation of the Human Cerebral Cortex from Intrinsic Functional Connectivity MRI. *Cereb Cortex*. Published online 2017:1-20. doi:10.1093/cercor/bhx179
4. Yeo BTT, Krienen FM, Sepulcre J, et al. The organization of the human cerebral cortex estimated by intrinsic functional connectivity. *J Neurophysiol*. 2011;106(3):1125-1165. doi:10.1152/jn.00338.2011
5. Van Essen DC, Glasser MF, Dierker DL, Harwell J, Coalson T. Parcellations and Hemispheric Asymmetries of Human Cerebral Cortex Analyzed on Surface-Based Atlases. *Cereb Cortex*. 2012;22(10):2241-2262. doi:10.1093/cercor/bhr291
6. Frazier JA, Chiu S, Breeze JL, et al. Structural brain magnetic resonance imaging of limbic and thalamic volumes in pediatric bipolar disorder. *Am J Psychiatry*. 2005;162(7):1256-1265. doi:10.1176/appi.ajp.162.7.1256
7. Brett M, Anton JL, Valabregue R, Poline JB. Region of interest analysis using an SPM toolbox. Poster presented at: 8th International Conferance on Functional Mapping of the Human Brain; June 2, 2002; Sendai, Japan. marsbar.sourceforge.net
8. Baranger DAA, Lindenmuth M, Nance M, et al. The longitudinal stability of fMRI activation during reward processing in adolescents and young adults. *NeuroImage*. 2021;232:117872. doi:10.1016/j.neuroimage.2021.117872
9. Poldrack RA, Mumford JA, Schonberg T, Kalar D, Barman B, Yarkoni T. Discovering Relations Between Mind, Brain, and Mental Disorders Using Topic Mapping. *PLOS Comput Biol*. 2012;8(10):e1002707. doi:10.1371/journal.pcbi.1002707
10. Yarkoni T, Poldrack RA, Nichols TE, Van Essen DC, Wager TD. Large-scale automated synthesis of human functional neuroimaging data. *Nat Methods*. 2011;8(8):665-670. doi:10.1038/nmeth.1635
11. Morgane PJ, Galler JR, Mokler DJ. A review of systems and networks of the limbic forebrain/limbic midbrain. *Prog Neurobiol*. 2005;75(2):143-160. doi:10.1016/j.pneurobio.2005.01.001
12. Green P, MacLeod CJ. SIMR: an R package for power analysis of generalized linear mixed models by simulation. *Methods Ecol Evol*. 2016;7(4):493-498. doi:10.1111/2041-210X.12504
13. Winkler AM, Ridgway GR, Douaud G, Nichols TE, Smith SM. Faster permutation inference in brain imaging. *NeuroImage*. 2016;141:502-516. doi:10.1016/j.neuroimage.2016.05.068
14. Simpson G. gavinsimpson/permute. Published online June 30, 2020. Accessed August 24, 2020. https://github.com/gavinsimpson/permute
15. Winkler AM, Webster MA, Vidaurre D, Nichols TE, Smith SM. Multi-level block permutation. *NeuroImage*. 2015;123:253-268. doi:10.1016/j.neuroimage.2015.05.092
16. Nakagawa S, Schielzeth H. Repeatability for Gaussian and non-Gaussian data: a practical guide for biologists. *Biol Rev*. 2010;85(4):935-956. doi:10.1111/j.1469-185X.2010.00141.x
17. Olvera Astivia OL, Gadermann A, Guhn M. The relationship between statistical power and predictor distribution in multilevel logistic regression: a simulation-based approach. *BMC Med Res Methodol*. 2019;19:97. doi:10.1186/s12874-019-0742-8
18. Forbes EE, Hariri AR, Martin SL, et al. Altered striatal activation predicting real-world positive affect in adolescent major depressive disorder. *Am J Psychiatry*. 2009;166(1):64-73. doi:10.1176/appi.ajp.2008.07081336

**Supplemental Table 1: Group comparisons of demographic characteristics and study procedures**

| | Lifetime History of Depression onset subgroups | | | |
|---|---|---|---|---|
| | **Early-onset (n=34)** | **Late-onset (n=24)** | ***t/χ²*** | ***p*** |
| Age in years (mean, SD) † | 18.44 (1.39) | 18.4 (1.39) | 0.1581 | 0.8746 |
| African American (N, percent) * | 17 (68%) | 27 (82%) | 0.8246 | 0.3638 |
| Financial disadvantage (N, percent) * | 16 (64%) | 14 (42%) | 1.8581 | 0.1728 |
| Pubertal tempo (mean, SD) ˣ | 0.35 (0.05) | 0.33 (0.05) | -1.0573 | 0.2954 |
| Pubertal timing (mean, SD,) ˣ | 10.23 (0.83) | 10.15 (0.94) | -0.3371 | 0.7373 |
| Number of scans (mean, SD,) ˣ | 2.6 (1.22) | 2.27 (1.01) | -1.086 | 0.2832 |
| Number of outlier frames (mean, SD) † | 22.8 (16.1) | 22.8 (20.7) | 0.416 | 0.679 |

Descriptives and comparisons of demographic variables between participants with early and late-onset MDD. †= group comparison was run as a linear mixed effect model controlling for participant ID as a random intercept. *= group comparison was run as a chi-squared test. ˣ= group comparison was run as a t-test.

**Supplemental Table 2: Age-related change in reward anticipation BOLD activity during late-adolescence, by network**

| **Network** | **Mean** | **SE** | **t** | **p** |
|---|---|---|---|---|
| Control | 0.25 | 0.09 | 2.60 | 0.01 |
| Default Mode | -0.03 | 0.06 | -0.43 | 0.67 |
| Dorsal Attention | 1.48 | 0.11 | 13.00 | $1.11 \times 10^{-16}$ |
| Limbic | -0.20 | 0.15 | -1.34 | 0.20 |
| Reward | 0.56 | 0.23 | 2.43 | 0.02 |
| Salience | 1.77 | 0.13 | 13.70 | $7.34 \times 10^{-18}$ |
| Somatomotor | 1.70 | 0.11 | 15.50 | $4.85x0^{-25}$ |
| Visual | 0.22 | 0.11 | 1.92 | 0.06 |

One-sample t-test testing whether the average age-related change in reward anticipation activation in each network is different from 0. Results are not corrected for spatial autocorrelation.

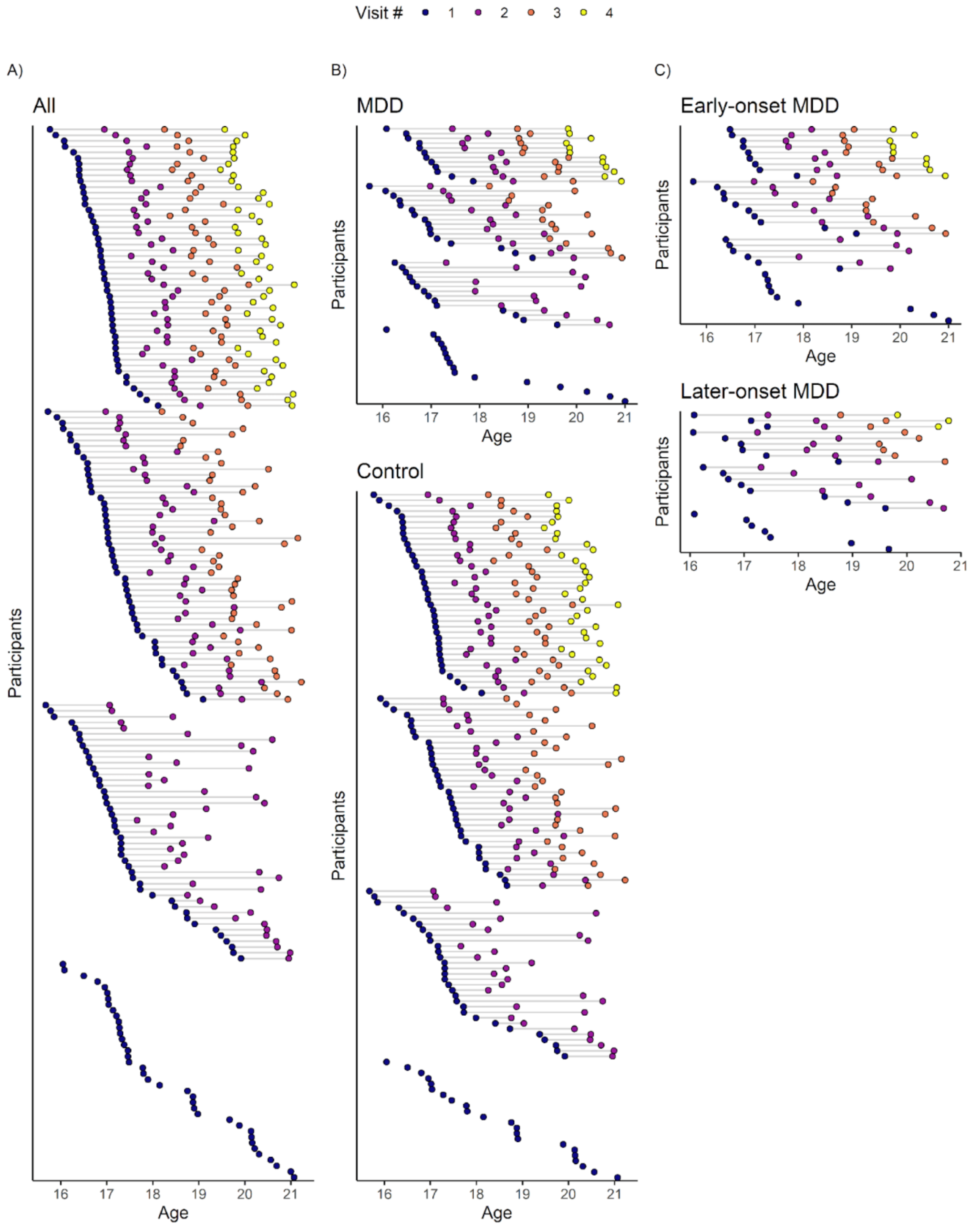


**Supplemental Figure 1:** Participant age at each study visit: Distribution of participant age, across and within waves, in the Pittsburgh Girls Study (PGS). MDD= Major Depressive Disorder. Control = No MDD.

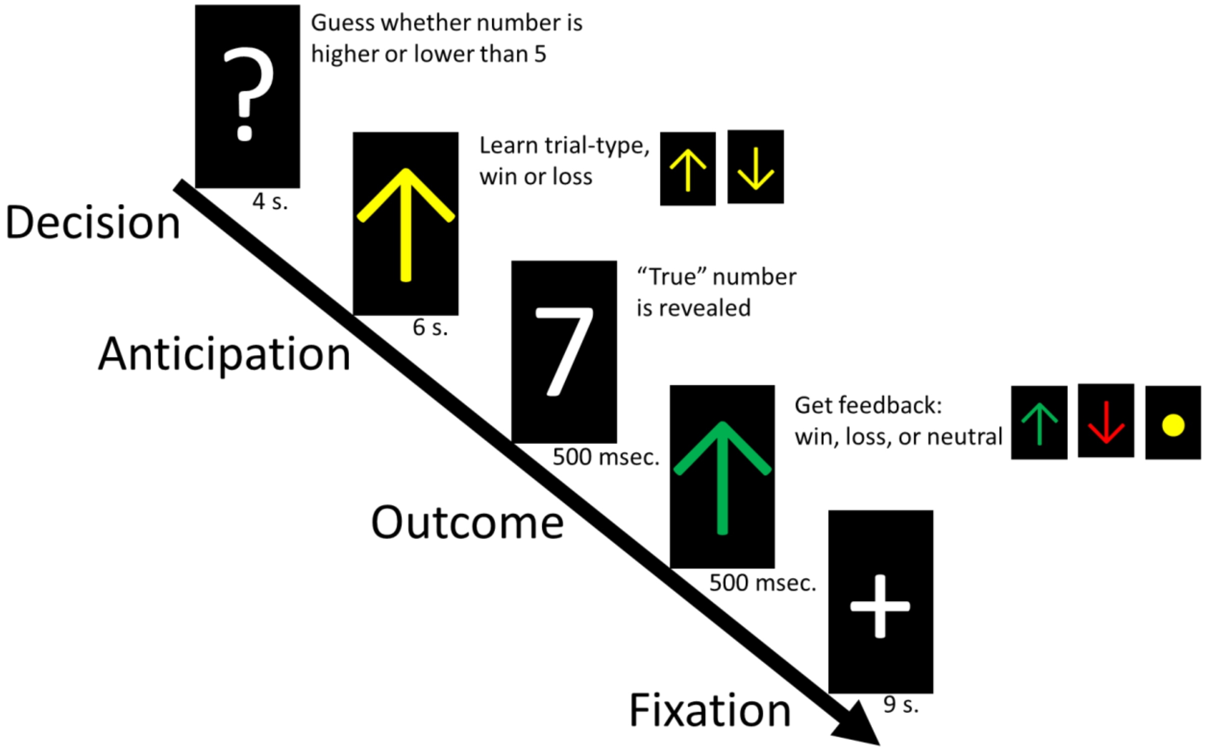


**Supplemental Figure 2:** Monetary Task design: Schematic representation of the fMRI task design, see Methods. After Forbes et al. [18]

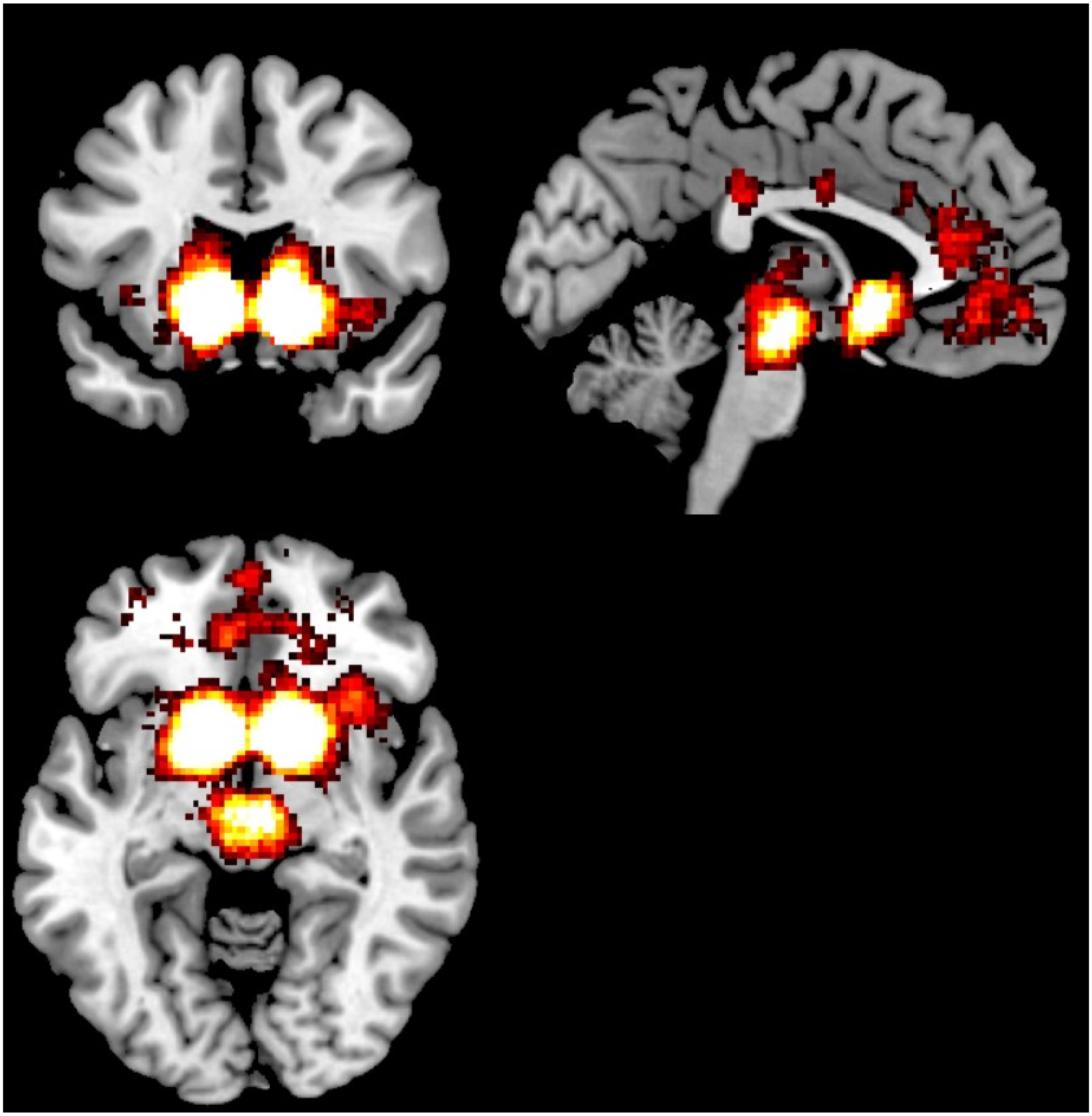

**Supplemental Figure 3:** Reward Network from Neurosynth: Results from Neurosynth [10] used to select regions to include in the 'Reward' network. Results were accessed from https://neurosynth.org/analyses/topics/v5-topics-50/7. Across 14,371 studies, the 1,218 reward-tagged studies are more likely to report significant activation in these regions than non-reward studies ($p<0.01$, FDR-corrected). Regions that likely to be reported in a 'reward' study include the caudate, putamen, nucleus accumbens, pallidum, amygdala, and thalamus, as well as portions of the orbitofrontal cortex, ventral medial prefrontal cortex, medial prefrontal cortex, insula, and the anterior cingulate.

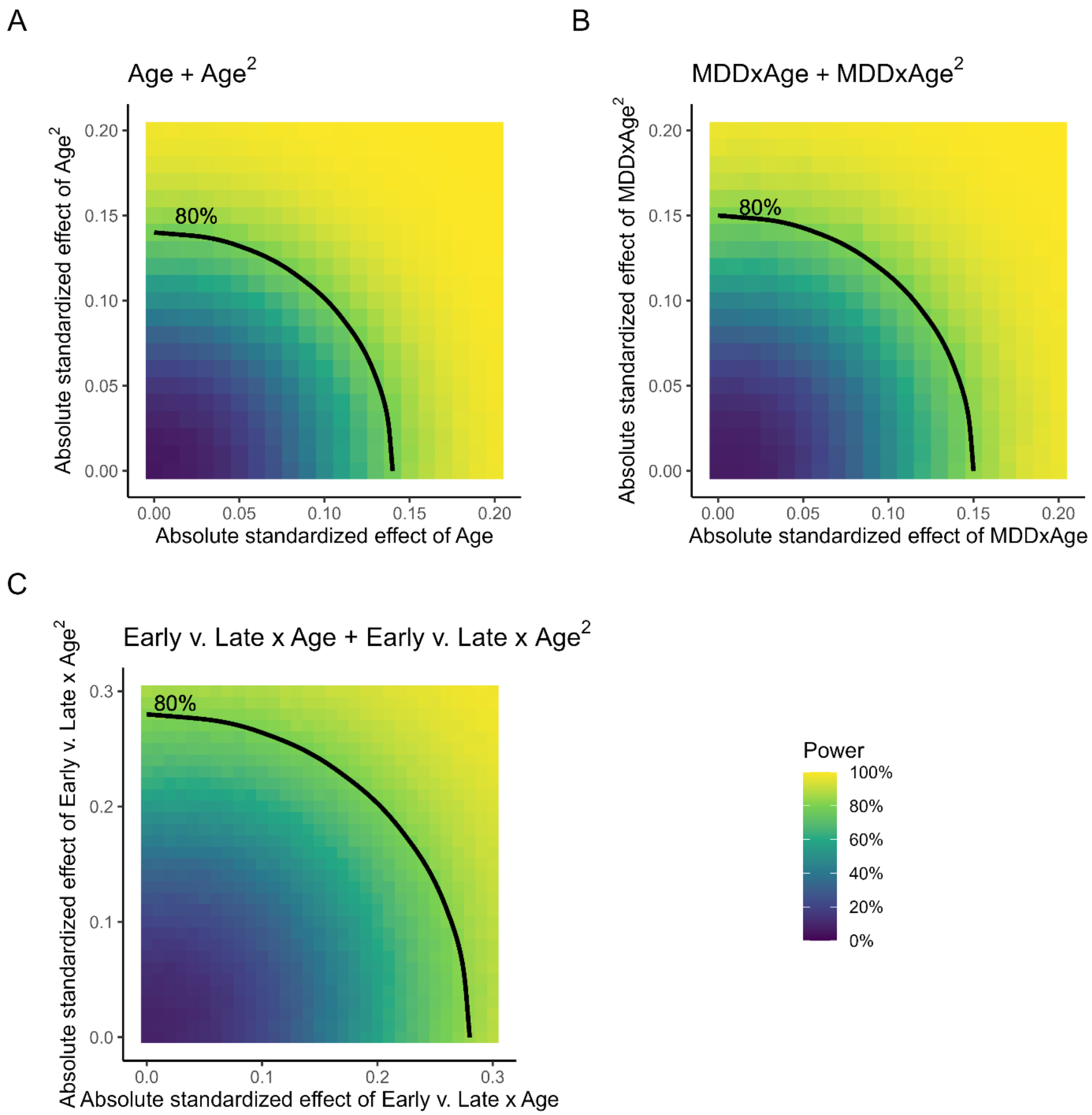


**Supplemental Figure 4.** Sensitivity analyses conducted using the R package simr. Plots are colored by power (the percent of 5,000 simulations where $p<0.05$) for each respective model. Curved lines demark the location of effects that achieve 80% power.

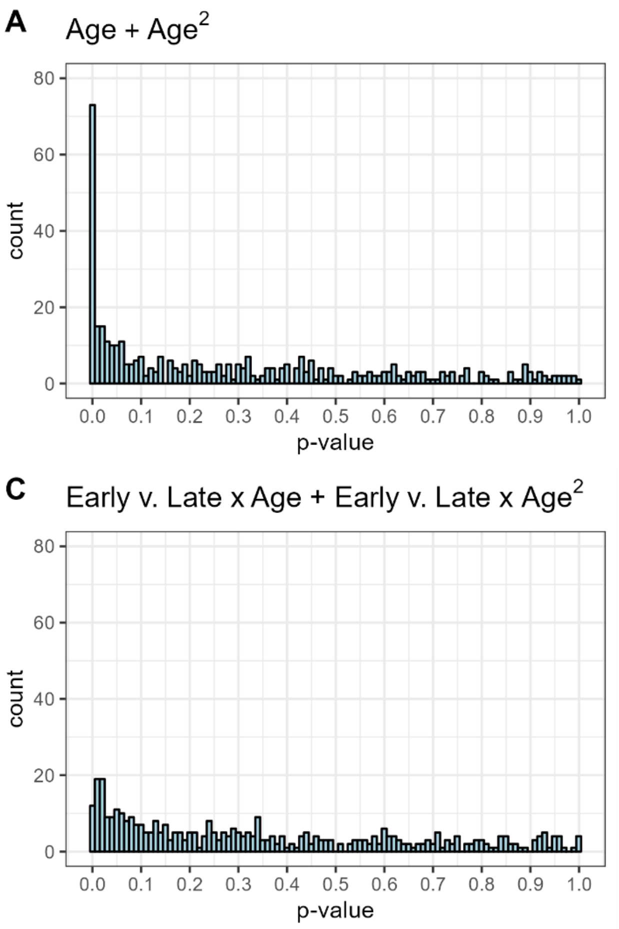


**Supplemental Figure 5.** Distribution of uncorrected p-values for primary analyses. The uniform distribution of null p-values (i.e., values on the right-hand side of each graph) indicate that analyses meet the exchangeability assumption of the FDR correction used.

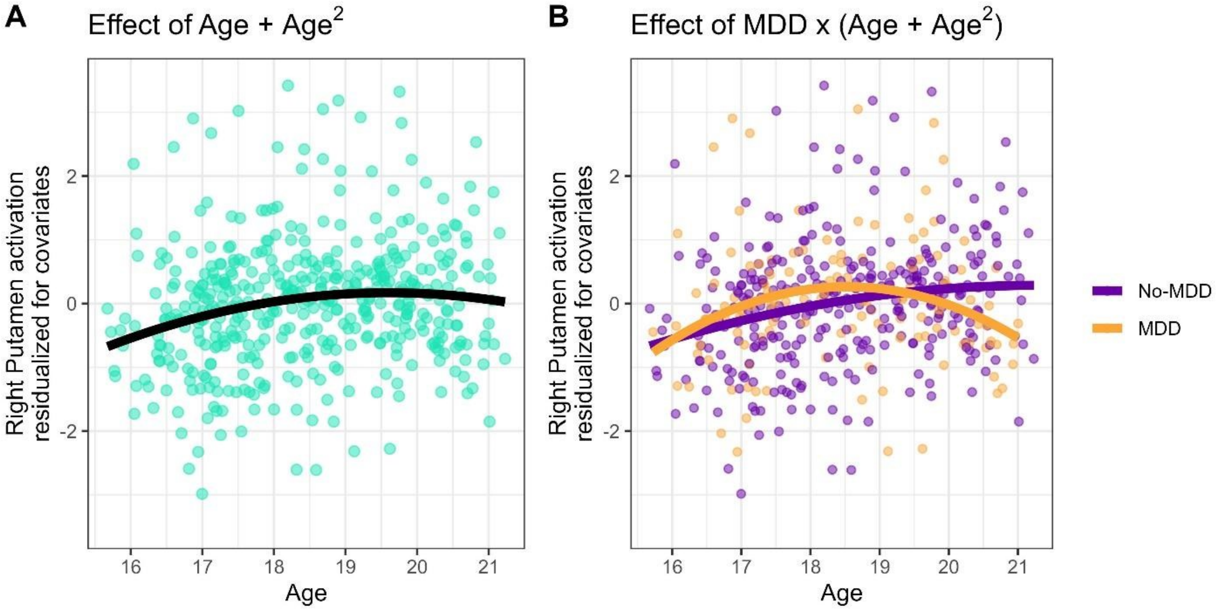


**Supplemental Figure 6.** Effects in the right putamen for Age (A) and MDD x Age (B). Activation was residualized for covariates (financial strain, pubertal tempo and timing, and number of outlier frames) in order to emphasize the effects of Age and MDD group. Both analyses are significant ($p<0.05$, fdr).

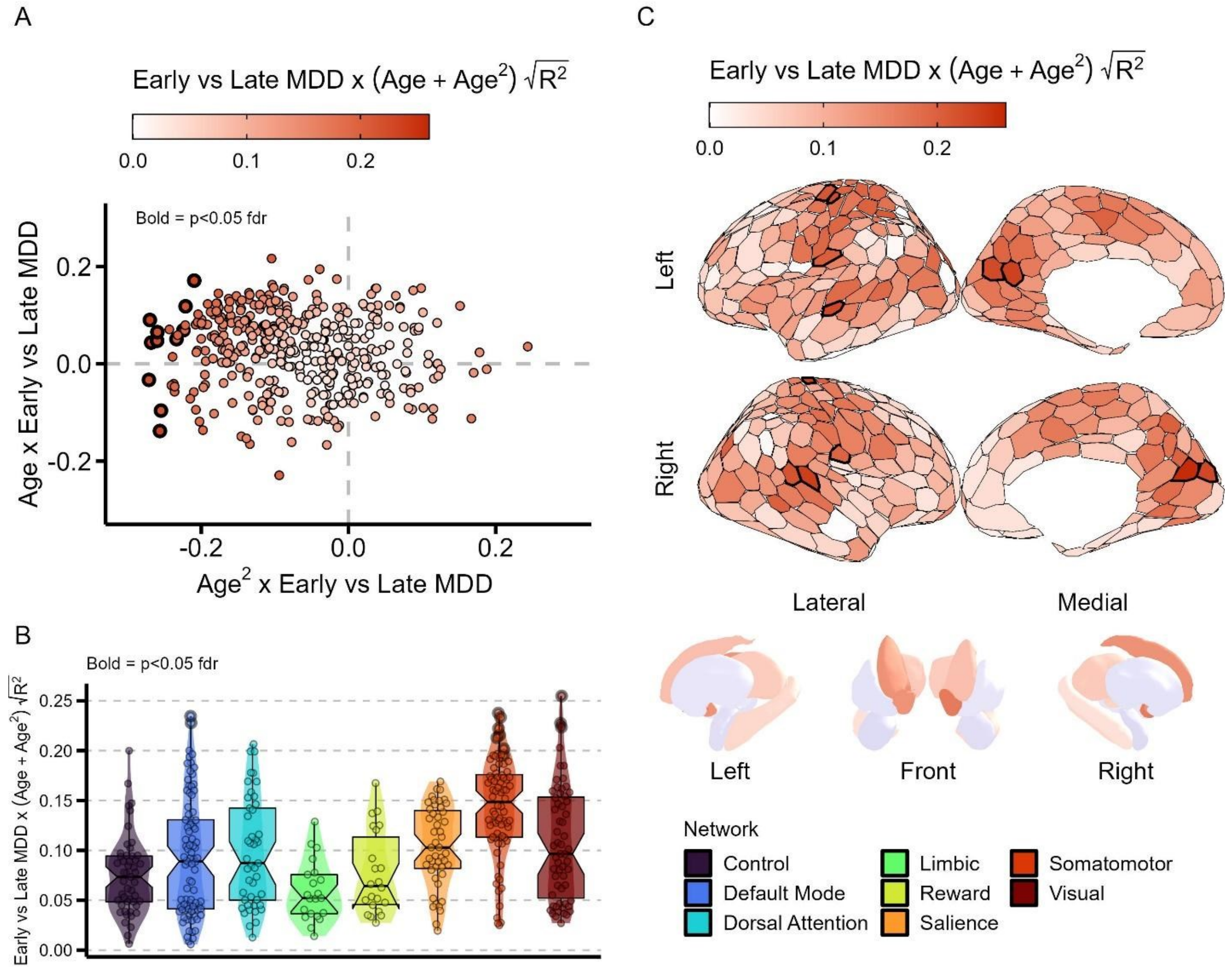


**Supplemental Figure 7. Greater adolescent developmental change in early-onset MDD**

**A**) Distribution of the interaction between age and $age^2$ with MDD-onset (early n=34; late n=24). Values are the standardized effect size of the interaction. Each point represents a separate region. Points are colored by the square-root of the increase in marginal $R^2$ from adding the Age x MDD-onset and $Age^2$ x MDD-onset interaction terms to the model, which is linearly proportional to the $\chi^2$ of the model comparison. Bolded points are significant p<0.05 fdr-corrected. **B**) The square-root of the marginal $R^2$ from age x MDD-onset and $age^2$ x MDD-onset interactions predicting reward activation, plotted by network. Significant (p<0.05 fdr) regions are bolded. Network-membership was not significantly associated with the MDD onset by age interaction effect ($F_{(7,401)}$=14.38, $p_{permutation}$=0.18). **C)** The square-root of the marginal $R^2$ from age x MDD-onset and $age^2$ x MDD-onset interactions predicting reward activation, plotted by region. Grey-blue regions and points reflect regions where the addition of the age x MDD-onset and $age^2$ x MDD-onset interactions worsened model fit. Significant (p<0.05 fdr) regions have a thicker outline.
